# Pharmacokinetics of nirmatrelvir and ritonavir in COVID-19 patients with end stage renal disease on intermittent haemodialysis

**DOI:** 10.1101/2022.08.19.22277959

**Authors:** Tilman Lingscheid, Martina Kinzig, Anne Krüger, Nils Müller, Georg Bölke, Pinkus Tober-Lau, Friederike Münn, Helene Kriedemann, Martin Witzenrath, Leif E. Sander, Fritz Sörgel, Florian Kurth

## Abstract

**Background:** Nirmatrelvir/ritonavir is an effective therapy against SARS-CoV-2. Patients with end-stage renal disease (ESRD) are at high risk for severe COVID-19 and show impaired vaccine responses underlining the importance of antiviral therapy. However, use of nirmatrelvir/ritonavir is not recommended in these patients due to lack of clinical and pharmacokinetic data.

**Objective:** To investigate pharmacokinetics and hepatic tolerance of nirmatrelvir/ritonavir in patients with ESRD and haemodialysis (HD).

**Patients and methods:** Four patients diagnosed with SARS-CoV-2 infection received nirmatrelvir/ritonavir 150/100mg twice daily as recommended for renal impairment; HD ran in two- to three-day intervals. Plasma and serum samples were drawn before and after each HD during the 5-day treatment and for ensuing 3-5 days.

**Results:** Median peak levels of nirmatrelvir obtained two hours after medication pre-HD in three patients were 7745ng/mL on day 3 and 6653ng/mL on day 5; median post-HD levels (C_6h_) declined to 5765ng/mL (74%) and 5521ng/mL (83%), on days 3 and 5 of treatment, respectively. Three days after end of treatment, median levels were 365ng/mL pre-HD and 30ng/mL post-HD. Measurements of the fourth patient, six hours after drug intake pre-HD showed nirmatrelvir-levels of 3704ng/mL on treatment day 3 which fell to 2308ng/mL post-HD, at one hour before intake of the next dose (C_min_).

**Conclusion:** Use of nirmatrelvir/ritonavir in patients with ESRD results in high nirmatrelvir blood concentrations, which are still within the range known from patients without renal failure. No accumulation of nirmatrelvir took place and levels declined to zero within few days after end of treatment.

## Introduction

Nirmatrelvir/ritonavir (N/r) has recently been introduced as antiviral therapy with high efficacy against SARS-CoV-2 and is approved in multiple countries for early treatment of high-risk patients with mild to moderate COVID-19.^1,2^ End stage renal disease (ESRD) is a major risk factor for severe COVID-19 with high prevalence in many western populations. Moreover, immunogenicity and effectiveness of COVID-19 vaccines is reduced in patients with ESRD,^3^ making them an important target group for additional, preventive treatment.

Nirmatrelvir is rapidly metabolized via CYP3A4 when given alone. In combination with ritonavir, almost no metabolization of nirmatrelvir is observed and the drug is excreted primarily through renal elimination.^4^ In patients with moderate renal impairment (estimated glomerular filtration rate [eGFR] 31-59mL/min) dose adjustment is recommended from 300mg/100 mg BID to 150mg/100 mg BID, in order to avoid accumulation. In patients with severe renal impairment (eGFR <30mL/min) including ESRD and haemodialysis (HD), use of N/r is currently not recommended due to insufficient pharmacokinetic and clinical data.^5-7^ Dose adjustments have been proposed and a recent case series investigated a dose of 300/100 mg N/r on day one, followed by 150/100 mg once daily from day two to five in ESRD patients, yet, without pharmacokinetic measurements.^8,9^

Additional assessment of the pharmacokinetic behaviour of N/r in patients with ESRD on chronic intermittent HD is therefore of particular interest for clinicians, especially because of the increased risk of a severe course of COVID-19 in these patients.^7,10,11^ Herein we report data on pharmacokinetics and hepatic tolerance of nirmatrelvir and ritonavir treatment in four ESRD patients on intermittent HD with COVID-19.

## Methods

### Patients / treatment / medication

Four patients undergoing outpatient HD therapy at our centre tested positive for SARS-CoV-2 in 2022. Based on the individual risk profile of the patients, the nephrologists in charge initiated antiviral therapy; treatment with N/r was chosen because of the possibility of oral administration without need for hospital admission or hospital visits in addition to the outpatient HD sessions.

Treatment was started three to five days after the first positive SARS-CoV-2 RT-PCR test. All patients received the recommended reduced dose for renal impairment of 150 mg nirmatrelvir and ritonavir 100 mg BID orally at 7 AM and 7 PM for five days; patient #1 was on constant ritonavir therapy (100 mg OD), due to a well-controlled HIV-infection and continued this medication after finishing the 5-day course of N/r BID. Patient characteristics including concomitant medication are shown in the supplementary material. In patients #1, #2 and #3 HD started in the morning 2-3 hours after intake of the medication. In patient #4, HD started approximately 6 hours after the morning dose.

### Haemodialysis and Bio-sampling

Blood-sampling was performed as part of clinical routine within the framework of an observational study rather than sampling within an interventional trial. HD was performed using the Artis Physio® (Baxter Deutschland GmbH, Unterschleißheim) dialysis machine with a high-flux dialyzer (Revaclear 400 ®, Baxter). Duration of each dialysis session was 240 minutes for patients #1, #2 and #3; 300 minutes for patient #4. In all cases venous access was established via double AV-fistula puncture. Blood-flow rates were 250-300 mL/min with dialysate flow rates of 500 mL/min for all patients. Net ultrafiltration rates of 500-600 mL/h for patient #1, 900-1100mL/h for patient #2, 750-1200mL/h for patient #3 and 800-900mL/h for patient #4 were achieved. Unfractionated heparin was given to patient #1, fractioned heparin to patient #2, whereas in patients #3 and #4 a citrate anticoagulation protocol was used. Blood samples (plasma and serum) were drawn at the beginning and the end of each HD session for each patient (except first sample of patient #1: only serum sample). Samples of post-filter dialysate were drawn at 0.5, 2 and 4 hours after start of HD. Further information is given in the supplementary material.

### Determination of nirmatrelvir/ritonavir levels

N/r concentrations were quantified in human plasma, serum and dialysate by liquid chromatography with tandem mass spectrometry (LC-MS/MS) using an API 5500 triple quadrupole mass spectrometer (SCIEX, Concord, Ontario, Canada). A volume of 25 µL of each sample was deproteinized with 150 µL acetonitrile (containing the internal standard piperacillin-d5), subsequently vortex-shaken and centrifuged. The supernatant was further diluted with Milli-Q® water and 10 µL of each sample was injected into the LC-MS/MS system. Quantification of N/r was performed by peak area ratio of analytes to internal standard.

The linearity of the calibration curve for nirmatrelvir was proven from 14.69 to 4753 ng/mL and for ritonavir from 10.87 to 3516 ng/mL. The lower limit of quantification in all matrices was set to 14.69 ng/mL for nirmatrelvir and to 10.87 ng/mL for ritonavir. No interferences in the chromatogram were observed for nirmatrelvir and ritonavir and the internal standard in human plasma, serum and liquor. Intra-and inter-day precision of the spiked quality control samples was below 5% with an intra-and inter-day accuracy between 90% and 110%.

### Data

Data was analysed and visualized with Graphpad Prism Version 9; aggregated data is summarized as median and interquartile range.

### Ethics

The described patients are part of the Pa-COVID-19 cohort study, a prospective observational study conducted at the tertiary care university hospital of Charité – Universitätsmedizin Berlin.^12^ Written informed consent was provided by all patients prior to inclusion. The study was approved by the ethics committee of Charité – Universitätsmedizin Berlin (EA2/066/20), is conducted according to the Declaration of Helsinki and Good Clinical Practice principles (ICH 1996) and registered in the German and WHO international clinical trials registry (DRKS00021688).

## Results

Treatment courses, HD sessions and pharmacokinetic measurements of nirmatrelvir and ritonavir in plasma and dialysate of individual patients are shown in **Figure 1**.

**Figure 1:**
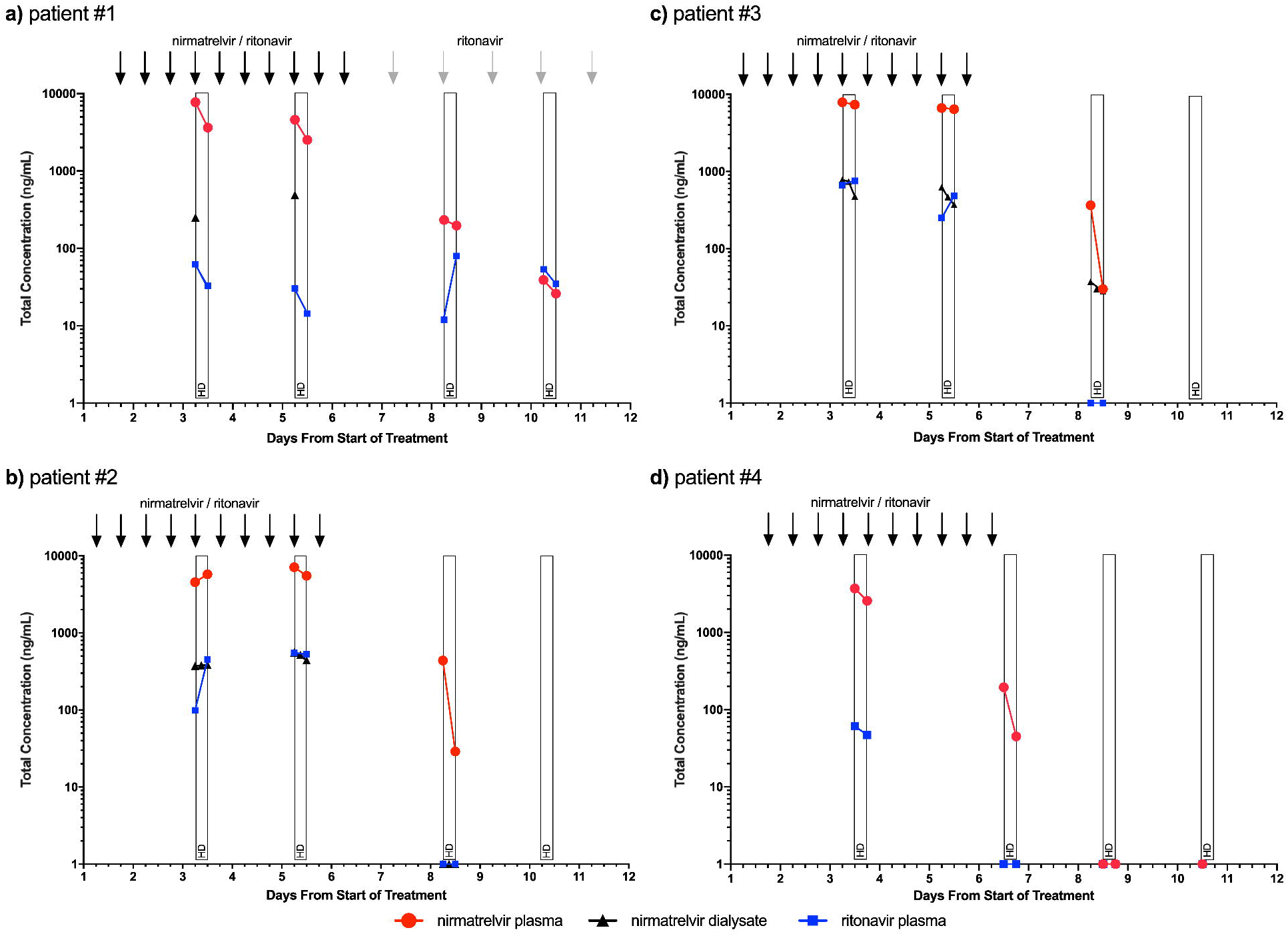
**A:** patient #1, **B:** patient #2. **C**: patient #3, **D**: patient #4: pharmacokinetics of nirmatrelvir in plasma (red circles) and dialysate (black triangles) and ritonavir in plasma (blue squares) sampled pre- and post-haemodialysis (HD) during and after the five-day course of nirmatrelvir/ritonavir (N/r, days 1-5). Concentrations the limit of quantification are positioned on the x-axis not suggesting that there is a measurable level but in order to demonstrate zero results on the logarithmic scale of the y-axis. Each thick black arrow resembles one dose of N/r (150/100 mg); gray arrows represent one dose of ritonavir (100 mg) in patient #1. Pre-HD sample of patient #1 on day 3 is a serum sample instead of plasma.

Patient #1 presented with a peak nirmatrelvir concentration of 7745 ng/mL two hours after intake of his fourth dose of 150 mg/100 mg N/r. During the four-hour HD course, plasma concentration fell by 53% to 3636 ng/mL. At the second sampling on day five a peak plasma concentration of 4601 ng/mL was measured which fell by 45%, to 2518 ng/mL post-HD. Consecutive measurement three days later exhibited concentrations around 200 ng/mL, most likely due to the continued ritonavir intake; detailed individual plasma measurements are shown in **Table A1**; ritonavir concentrations in plasma ranged between 14 and 120 ng/mL (**Table A3**). Patients #2 and #3 both received HD on treatment day three after intake of the fifth dose of N/r. Nirmatrelvir levels of patient #2 increased during HD from 4563 ng/mL to 5765 ng/mL; Patient #3 showed stable concentrations of 7898 ng/mL and 7345 ng/mL pre- and post-HD, respectively (7% decrease). Plasma levels on treatment day 5 were 7116 ng/mL pre-HD and 5521ng/mL post-HD in patient #2 and 6653 ng/mL pre-HD and 6417 ng/mL post-HD in patient #3. Measurements on day 8 (3 days after end of treatment) showed levels of 438 and 365 ng/mL pre-HD with a decline to 29 and 30 ng/mL, in patients #2 and #3, respectively. Ritonavir levels were comparatively high, ranging from 99 to 553 ng/mL in patient #2 and from 250 to 756 ng/mL in patient #3.

Patient #4 presented with a nirmatrelvir plasma concentration of 3704 ng/mL six hours after intake of the fourth dose of 150 mg/100 mg N/r. Post-HD plasma concentration fell by 38% to 2308 ng/mL at 11-12 hours after drug intake. Measurement after the last dose on day six showed concentrations of 187 ng/mL and 47 ng/mL pre- and post-HD, respectively. Further measurement showed nirmatrelvir levels below the limit of quantification, likewise to negative ritonavir levels (**Figure 1, Table A1** and **A3**).

According to the known pharmacokinetic profiles of nirmatrelvir in healthy volunteers,^7^ we regarded the plasma concentrations in patients #1-3, measured at 2-3 hours after drug intake, as maximum plasma concentrations (C_max_). These were in median 7745 ng/mL on treatment day three, declining to a median of 5765 ng/mL post-HD. Patient #1 had taken four doses of N/r at this time point, patients #2 and #3 had taken 5 doses due to different starting time points of therapy. Median C_max_ levels on treatment day 5 were 6653 ng/mL, declining to 5521 ng/mL post-HD. On day eight (three days after end of treatment) median levels were 365 and 30 ng/mL pre- and post-HD, respectively (**Figure 2)**.

**Figure 2:**
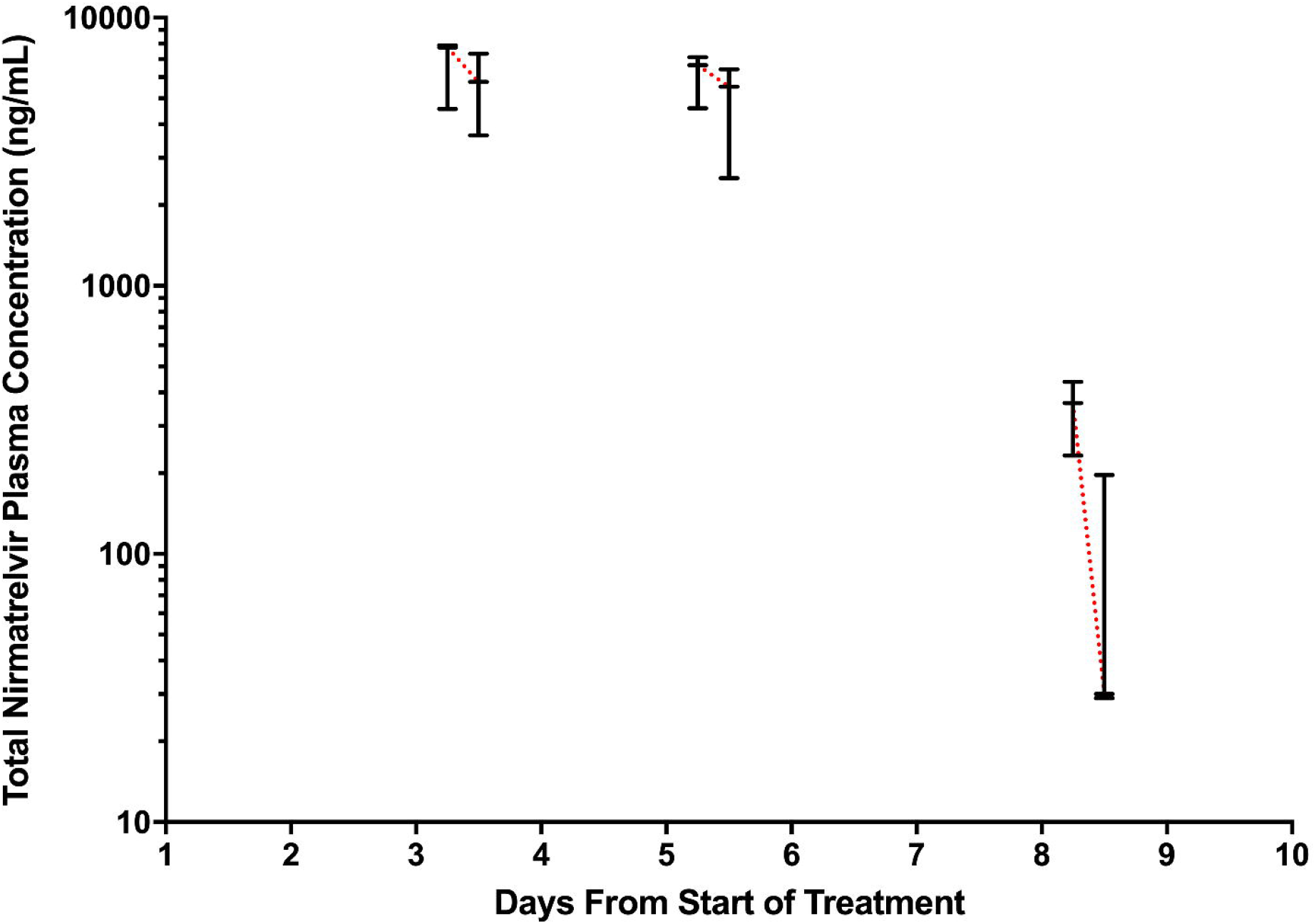
Pooled data of nirmatrelvir concentrations pre- (C_2h/max_) and post-haemodialysis (C_6h_) of patients #1-3 on treatment days 3, 5 and 8 (i.e. 3 days post treatment); connected centred bars depict the median, bars depict the range.

Levels of nirmatrelvir in dialysate were measured in patient #1 during the first and second HD session at 30 min after HD start: levels of 254 ng/mL and 489 ng/mL were detected. Patients #2 and #3 received full dialysate sampling with data shown in **Figure 1** and **Table A2**. No dialysate sampling was done in patient #4. In general, concentrations measured in dialysate corresponded to the plasma levels in individual patients.

Nirmatrelvir blood concentrations were measured in plasma and serum for evaluation and alignment of both collection types regarding measurement accuracy. Measurements are shown in **Table A1**. Mean accuracy varied between 83 and 109%.

Ritonavir plasma levels varied to a very high extent between the different patients with measurements ranging from 14 ng/mL to 756 ng/mL (**Table A3)**. Measurements of ritonavir in plasma and serum are shown in **Table A3**; accuracy varied between 86 and 113%.

Overall, no signs of relevant drug-related toxicity or hepatic impairment were observed (**Figure A1**). An isolated and pre-existing increase of alkaline phosphatase among all patients remained unchanged or resolved spontaneously after end of treatment. No patient reported severe adverse events attributed to the medication.

All patients had a moderate course of COVID-19; patient #1 was tested negative for SARS-CoV-2 on day 10 after diagnosis. Patient #2 had a low viral load seven days after initial diagnosis and patient #3 after ten days. Patient #4 remained SARS-CoV-2 positive for six weeks. Initial viral load from nasopharyngeal swabs was high (ct-value <30), initially decreased (ct-value >30), spiked again 5 days after end of treatment and then remained positive with a low viral load for further four weeks

## Discussion

We report pharmacokinetic data of N/r in four patients with ESRD and intermittent HD. To our knowledge, this is the first data on pharmacokinetics of N/r in this group of patients, who constitute an important high-risk population for severe COVID-19. Existing pharmacokinetic data on nirmatrelvir are mainly available from assessment reports such as the European Medicines Agency’s (EMA) report and data from the US Food and Drug Administration (FDA).^5,7^

Measured peak plasma concentrations of nirmatrelvir were roughly within the 90% prediction intervals for nirmatrelvir at steady state in patients with normal kidney function.^5,7^ The 95^th^ predicted nirmatrelvir C_max_ on day five of treatment is assumed to be 10000 ng/mL among patients without renal impairment;^5^ this level was not reached at peak measurements 2-3 hours after drug intake in our patients despite ESRD and twice daily medication of nirmatrelvir. Peak levels were hence slightly higher than those measured on average in healthy volunteers and in patients without renal impairment, yet, no accumulation in general of nirmatrelvir was observed.

Exact C_min_ levels were not determined in our study as HD sessions of the majority of patients ran early in the morning soon after drug intake; the post-HD sample of patient #4, however, corresponds well to C_min_ as it was sampled approximately 11 hours after drug intake, one hour before intake of the evening dose of N/r. The respective nirmatrelvir concentration of 2308ng/mL, again, is within the 90% prediction interval at steady state, yet, clearly above the IC_90_ of 292 ng/mL of nirmatrelvir.^5,7^ The recently proposed dose reduction of N/r to 150/100 mg once daily after initial loading with 300/100 mg may reduce high peak levels but may still maintain C_min_ levels above the IC_90_ considering the elimination of nirmatrelvir from plasma observed in our case series.^5^

After the end of treatment, nirmatrelvir levels declined rapidly but were still measurable three days after treatment, whereas ritonavir levels were not measurable. Nirmatrelvir plasma levels pre-HD on day eight ranged from 233 to 438 ng/mL among patients #1-3; still partly reaching the IC_90_ of 292 ng/mL for SARS-CoV-2.^5,7^ Thus, antiviral exposition to nirmatrelvir may be longer in patients with ESRD, which may be a welcome effect in light of the debate of relapses after N/r therapy and proposed longer treatment phases.^13^ On the contrary, patient #4 had sub-therapeutic nirmatrelvir levels on day four of treatment. In addition, ritonavir-levels were below the detection limit as well; the patient ensured having taken the whole course of N/r. Possible explanation for the low concentrations measured on day four in this patient include individual problems with drug intake or absorption in this patient or highly variable elimination.

Ritonavir plasma levels were highly variable but in general within the range of earlier published data. However, data on ritonavir plasma levels are scarce and have to be interpreted with care.^14,15^ Nevertheless, the high nirmatrelvir levels indicate adequate ritonavir exposure in spite of variable plasma concentrations.

In summary, our data suggests that use of N/r in patients with ESRD and HD results in peak and minimum blood concentrations at the higher end of levels observed in patients without ESRD. However, no accumulation of nirmatrelvir was observed and plasma levels declined rapidly within a few days after end of treatment.

## Data Availability

All data produced in the present study are available upon reasonable request to the authors

## Transparency declarations

None to declare.

## Acknowledgements

We thank all members of the Pa-COVID-19 collaborative study group, and the dialysis team.

