## Supplementary Material for "Pharmacokinetics of nirmatrelvir and ritonavir in COVID-19 patients with end stage renal disease on intermittent haemodialysis"

**Patients:**

**Patient #1**: age: late sixties; male.

Relevant medical history: diabetic nephropathy, intermittent haemodialysis for four years, no residual renal function; diabetes mellitus type 2, COPD GOLD IV, peripheral arterial disease, coronary artery disease, HIV since 10 years.

Relevant medication: aspirin 100mg od, insulin varying doses, fluvastatin 40mg od (paused), clopidogrel 75mg od (paused), telmisartan 40mg od, cinacalcet 30mg od, raltegravir 400mg bid, lamivudine 100mg od, darunavir 800mg od, ritonavir 100mg od.

**Patient #2**: age: early sixties; male.

Relevant medical history: hypertensive nephropathy, intermittent haemodialysis for three years, no residual renal function coronary artery disease, peripheral arterial disease diabetes mellitus type 2.

Relevant medication: aspirin 100mg od, atorvastatin 40mg od paused, l-thyroxin 50µg od, pantoprazole 40mg od, tilidin 100mg bd, varying insulin doses,

**Patient #3**: age: late seventies; male.

Relevant medical history: diabetic nephropathy, intermittent haemodialysis for two years, no residual renal function, liver cirrhosis CHILD B, diabetes mellitus type 2, coronary artery disease, atrial fibrillation.

Relevant medication: aspirin 100mg od, simvastatin 20mg od (paused), pantoprazole 20mg od, pregabalin 25mg od, semaglutide 1mg twice weekly, insulin varying doses, allopurinol 100mg od.

**Patient #4**: age: late sixties; male.

Relevant medical history: diabetic/hypertensive nephropathy, intermittent haemodialysis for two years, no residual renal function, diabetes mellitus type 2, coronary artery disease, atrial fibrillation.

Relevant medication: bisoprolol 5mg bid, insulin varying doses, pantoprazole 20mg bid, clopidogrel 75mg od (paused), pregabalin 25mg od, sevelamer 800mg tid.

**Additional Methods:**

Sampling: K3EDTA liquid blood collection tubes containing tri potassium ethylene diamine tetra acetic acid to prevent clotting and CAT serum separator tubes (Greiner Bio-One, Austria) were used and processed within a maximum of one hour. Plasma/serum was separated by centrifugation and stored at -80°C prior to pharmacokinetic analyses. Dialysate was collected in no additive tubes (Greiner Bio-One, Austria) aliquoted and stored likewise.

**Figure A1**: Monitoring of Liver Functioning Tests on Haemodialysis Days.

A: Patient #1

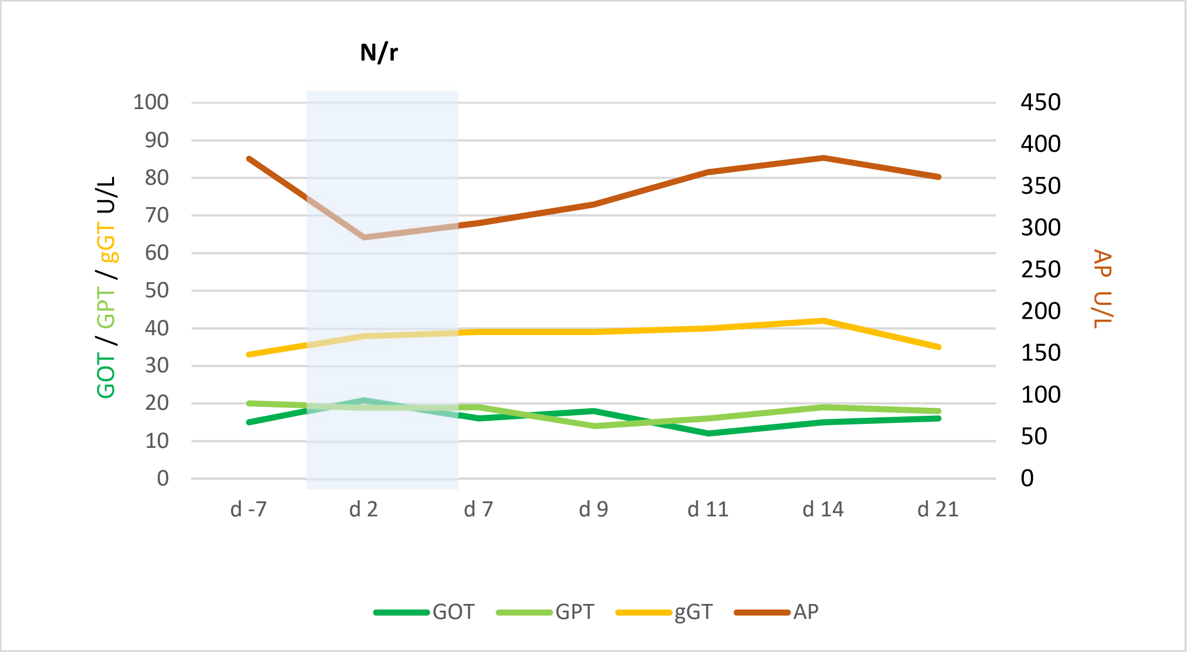


B: Patient #2

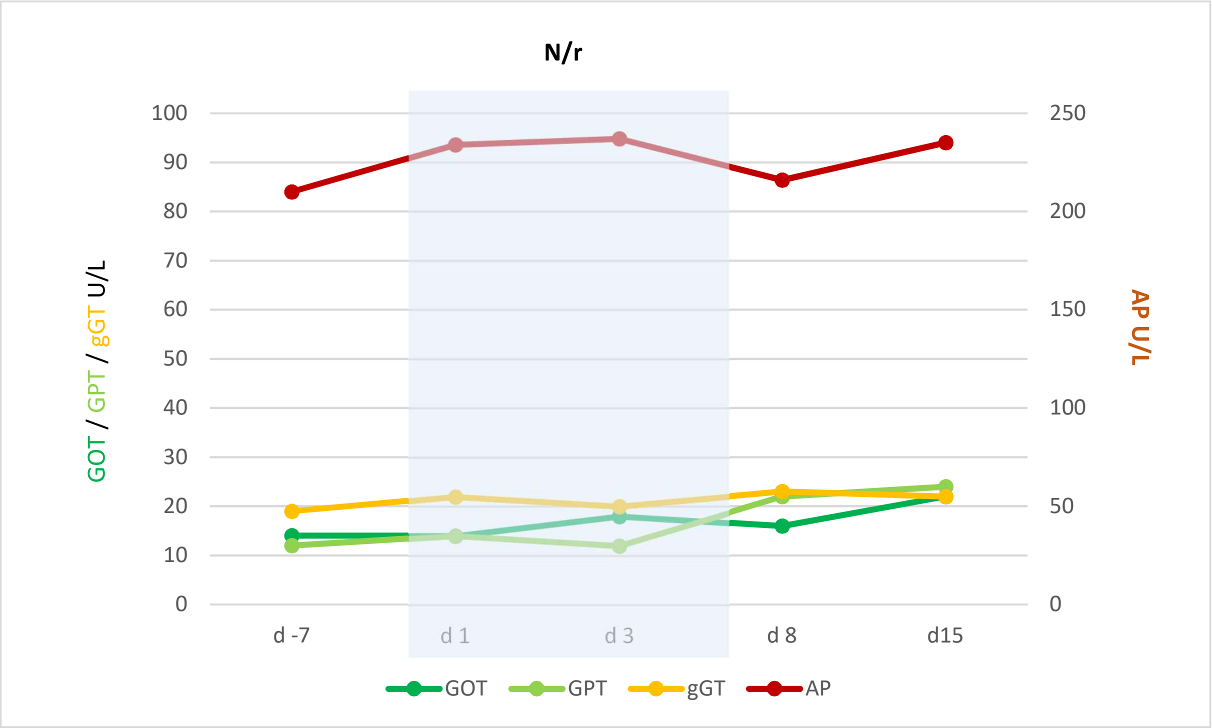

C: Patient #3

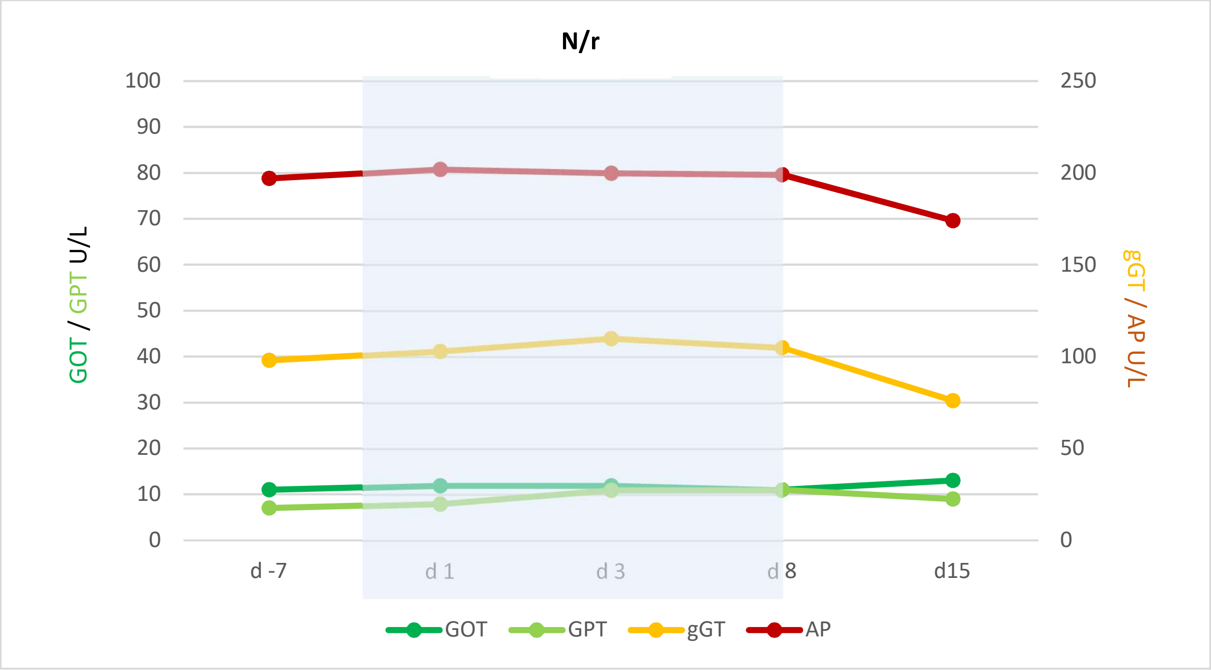


D: Patient #4

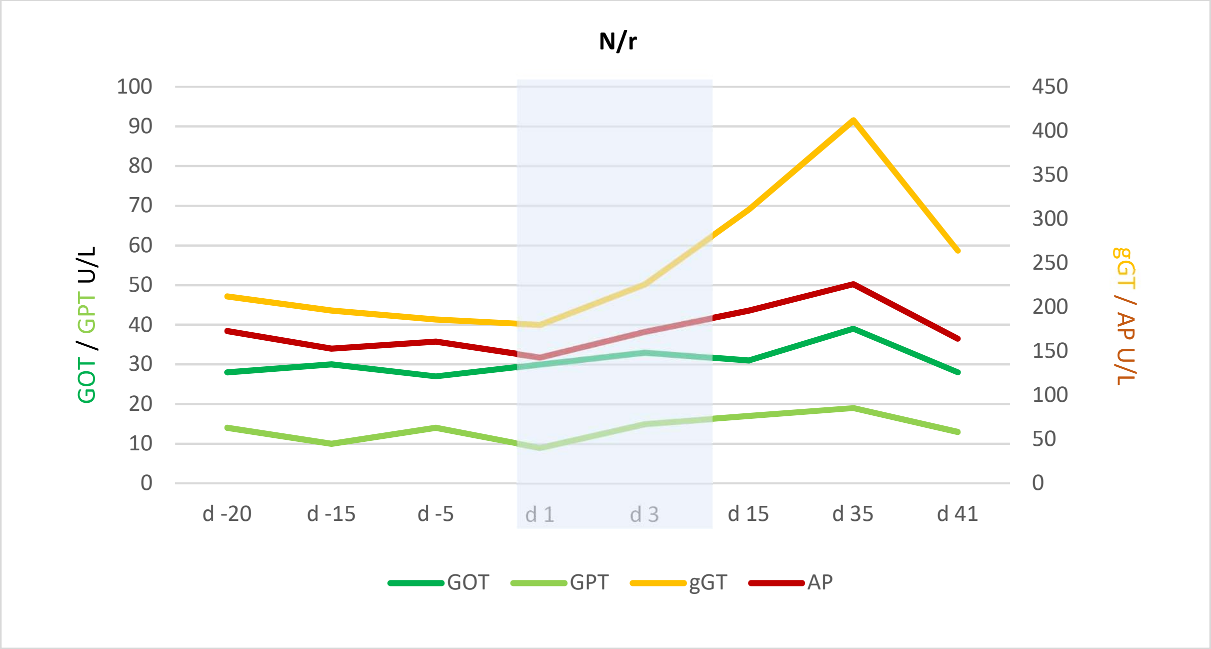


GOT: glutamic-oxaloacetic transaminase (upper limit of normal (ULN): <50 U/L); GPT: glutamic pyruvic transaminase (ULN: <41 U/L); GGT: Gamma-glutamyltransferase (ULN: <61 U/L); AP: alkaline phosphatase (ULN: <130 U/L). Grey area depicts interval of treatment with N/r (nirmatrelvir/ritonavir)

**Table A1: Comparison of nirmatrelvir concentrations measured from a blood sample and either centrifuged as serum or EDTA plasma**

| Timepoint (days of treatment, start=D1) | Concentration Nirmatrelvir ng/mL in Plasma | Concentration Nirmatrelvir ng/mL in Serum | Accuracy (%) Plasma / Serum |
|---|---|---|---|
| **A: Patient #1** | | | |
| D3 pre-HD | - | 7745 | - |
| D3 post-HD | 3636 | 3826 | 95.0 |
| D5 pre-HD | 4601 | 4826 | 95.3 |
| D5 post-HD | 2518 | 2729 | 92.3 |
| D8 pre-HD | 232.8 | 255.8 | 91.0 |
| D8 post-HD | 197.2 | 203.2 | 97.0 |
| D10 pre-HD | 39.80 | 38.58 | 103.2 |
| D10 post-HD | 26.29 | 31.2 | 84.3 |
| D12 pre-HD | <LLOQ | <LLOQ | - |
| D12 post-HD | <LLOQ | <LLOQ | - |
| **B: Patient #2** | | | |
| D3 pre-HD | 4563 | 4560 | 100.1 |
| D3 post-HD | 5765 | 5314 | 108.5 |
| D5 pre-HD | 7116 | 7150 | 99.5 |
| D5 post-HD | 5521 | 6630 | 83.3 |
| D8 pre-HD | 437.8 | 437.8 | 100.0 |
| D8 post-HD | 29.04 | 29.06 | 99.9 |
| **C: Patient #3** | | | |
| D3 pre-HD | 7898 | 8596 | 91.9 |
| D3 post-HD | 7345 | 7411 | 99.1 |
| D5 pre-HD | 6653 | 6675 | 99.7 |
| D5 post-HD | 6417 | 6086 | 105.4 |
| D8 pre-HD | 364.7 | 342.7 | 106.4 |
| D8 post-HD | 29.97 | 30.47 | 98.4 |
| **D: Patient #4** | | | |
| D3 pre-HD | 3704 | 3778 | 98,0 |
| D3 post-HD | 2308 | 2572 | 89,8 |
| D6 pre-HD | 187.3 | 195.2 | 96,0 |
| D6 post-HD | 46.91 | 44.97 | 104,3 |
| D8 pre-HD | <LLOQ | <LLOQ | - |
| D8 post-HD | <LLOQ | <LLOQ | - |
| D10 pre-HD | <LLOQ | <LLOQ | - |

lower limit of quantitation (LLOQ)

### Table A2: measurements dialysate of nirmatrelvir and ritonavir

| Timepoint (days of treatment, start=D1) | Concentration Nirmatrelvir ng/mL in Dialysate | Concentration Ritonavir ng/mL in Dialysate |
|---|---|---|
| **A: Patient #1** | | |
| D3 30min HD | 254.0 | <LLOQ |
| D5 30min HD | 489.0 | <LLOQ |
| **B: Patient #2** | | |
| D3 30min | 372.5 | <LLOQ |
| D3 2h | 380.0 | <LLOQ |
| D3 4h | 384.3 | <LLOQ |
| D5 30min | 554.6 | <LLOQ |
| D5 2h | 519.4 | <LLOQ |
| D5 4h | 442.7 | <LLOQ |
| D8 30min | <LLOQ | <LLOQ |
| D8 2h | <LLOQ | <LLOQ |
| D8 4h | <LLOQ | <LLOQ |
| **C: Patient #3** | | |
| D3 30min | 783.7 | <LLOQ |
| D3 2h | 737.8 | <LLOQ |
| D3 4h | 475.4 | <LLOQ |
| D5 30min | 622.4 | <LLOQ |
| D5 2h | 466.1 | <LLOQ |
| D5 4h | 370.9 | <LLOQ |
| D8 30min | 37.3 | <LLOQ |
| D8 2h | 29.86 | <LLOQ |
| D8 4h | 28.08 | <LLOQ |

lower limit of quantitation (LLOQ)

**Table A3: Comparison of ritonavir concentrations measured from a blood sample and either centrifuged as serum or EDTA plasma**

| Timepoint (after treatment start = D1) | Concentration Ritonavir ng/mL in Plasma | Concentration Ritonavir ng/mL in Serum | Accuracy (%) Plasma / Serum |
|---|---|---|---|
| **A: Patient #1** | | | |
| D3 pre-HD | - | 62.3 | - |
| D3 post-HD | 33.1 | 35.1 | 94.2 |
| D5 pre-HD | 30.3 | 31.1 | 97.6 |
| D5 post-HD | 14.4 | 16.7 | 86.2 |
| D8 pre-HD | 120.2 | 123.2 | 97.6 |
| D8 post-HD | 79.7 | 83.8 | 95.1 |
| D10 pre-HD | 53.4 | 52.7 | 101.4 |
| D10 post-HD | 34.7 | 35.3 | 98.2 |
| D12 pre-HD | 72.0 | 76.8 | - |
| D12 post-HD | 51.1 | 52.3 | - |
| **B: Patient #2** | | | |
| D3 pre-HD | 98.72 | 115.3 | 85.6 |
| D3 post-HD | 451.9 | 446.7 | 101.2 |
| D5 pre-HD | 553.4 | 613.7 | 90.2 |
| D5 post-HD | 527.2 | 534.1 | 98.7 |
| D8 pre-HD | <LLOQ | <LLOQ | - |
| D8 post-HD | <LLOQ | <LLOQ | - |
| **C: Patient #3** | | | |
| D3 pre-HD | 667.5 | 617.9 | 108.0 |
| D3 post-HD | 756.1 | 667.2 | 113.3 |
| D5 pre-HD | 250.0 | 225.9 | 110.7 |
| D5 post-HD | 483.6 | 428.8 | 112.8 |
| D8 pre-HD | <LLOQ | <LLOQ | - |
| D8 post-HD | <LLOQ | <LLOQ | - |
| **D: Patient #4** | | | |
| D3 pre-HD | 63.4 | 61.0 | 103.8 |
| D3 post-HD | 46.9 | 48.6 | 96.4 |
| D6 pre-HD | <LLOQ | <LLOQ | - |
| D6 post-HD | <LLOQ | <LLOQ | - |
| D8 pre-HD | <LLOQ | <LLOQ | - |
| D8 post-HD | <LLOQ | <LLOQ | - |
| D10 pre-HD | <LLOQ | <LLOQ | - |

lower limit of quantitation (LLOQ)
